# The plasma proteome of plant-based diets: analyses of 2920 proteins in 49,615 people

**DOI:** 10.1101/2023.08.14.23294084

**Authors:** Tammy Y.N. Tong, Karl Smith-Byrne, Keren Papier, Joshua R. Atkins, Mahboubeh Parsaeian, Timothy J. Key, Ruth C. Travis

## Abstract

**Background & Aims:** Circulating proteins are integral to many biological processes and could be influenced by diet. We aimed to assess differences in the plasma proteome between people of different dietary groups, defined by degree of animal food consumption.

**Methods:** The UK Biobank recruited middle-aged adults (mostly 40 to 69 years) throughout the UK between 2006-2010. Relative concentrations of 2920 plasma proteins were quantified using the Olink Proximity Extension Assay on blood samples from 49,615 participants, who were also asked to report their ethnicity and consumption of red and processed meat, poultry, fish, dairy and eggs. We defined six diet groups among the white British participants (23,243 regular meat eaters, 23,472 low meat eaters, 486 poultry eaters, 1081 fish eaters, 721 vegetarians, and 54 vegans), and two diet groups among the British Indians (391 meat eaters and 167 vegetarians). We used multivariable-adjusted linear regressions to assess the cross-sectional differences in protein concentrations between diet groups, with correction for multiple testing.

**Results:** We observed significant differences in many plasma proteins by diet group (920 proteins in white British participants, 2 in British Indians). Of the biggest differences, compared with regular meat eaters, the non-meat eaters had significantly higher FGF21 (e.g. +0.40 SD in vegetarians), CKB (+0.34), GUCA2A (+0.33), FOLR1 (+0.32), IGFBP2 (+0.31) and DSG2 (+0.30); all groups except the vegans had lower HAVCR1 (-0.38 in vegetarians). Vegetarians also had significantly lower SELENOP (-0.46), while the vegans had lower FGFBP2 (-0.68). The observed differences were generally similar in direction in both ethnicities.

**Conclusions:** In this first comprehensive assessment of plasma proteins by diet group, we identified many differences in proteins between vegetarians, vegans and meat eaters; this variation in protein levels suggests differences in various biological activities, including gastrointestinal tract and kidney function, which may relate to differences in future disease risk.

## Introduction

Proteins are essential for many bodily functions including supporting cell and tissue growth and structural integrity, and for enabling a range of enzymatic, biochemical, signalling and transport functions across different systems in the human body. Proteomics describes the large-scale study of multiple proteins and their interconnected pathways, the investigation of which allows a more comprehensive understanding of disease mechanisms [1]. Protein expression can be altered by environmental factors, including diet [2]. Previous studies have shown that differences in dietary habits with varying levels of animal foods can substantially impact dietary intakes of protein, which supplies the nine essential amino acids that cannot be synthesised endogenously, as well as both dietary intakes and circulating concentrations of amino acids [3]. However, there is scant evidence on how diet may affect the proteome, partly due to the limited availability of proteomics data in large scale studies, and no prior studies have investigated how long-term vegetarian and vegan diets which limit or exclude animal food products may influence the proteome. Circulating proteins have an important role in the aetiology of multiple diseases [4,5], and hence the examination of dietary influences on the proteome may offer unique insights into understanding how vegetarian and vegan diets may affect future disease risk.

The aim of this study is to provide a detailed description of circulating protein concentrations in people with varying degrees of animal food consumption, using data from the UK Biobank.

## Methods

### Study Population

The UK Biobank is a prospective cohort study of around 500,000 middle-aged people (recruitment target 40-69 years), recruited from across the United Kingdom between 2006 and 2010. The scientific rationale and design of the UK Biobank have been described in detail previously [6]. In brief, participants were identified from National Health Service registers, and were invited to join the study if they lived within travelling distance (∼25 km) of one of the 22 assessment centres in England, Wales and Scotland. People who consented to participate in the study attended a baseline visit at the assessment centre where they completed a touchscreen questionnaire which asked about their lifestyle (including diet, alcohol consumption, smoking status, physical activity), socio-demographic characteristics and general health and medical history. All participants were also given a verbal interview, and had their physical measurements and blood samples taken by trained staff. The UK Biobank study was approved by the National Information Governance Board for Health and Social Care and the National Health Service Northwest Multicentre Research Ethics Committee (06/MRE08/65), and all participants gave informed consent to participate using a signature capture device at the baseline visit.

### Ethnicity classification

On the touchscreen questionnaire, participants were asked to self-identify their ethnicity as ‘White’, ‘Mixed’, ‘Asian or Asian British’, ‘Black or Black British’, ‘Chinese’, ‘Other ethnic group’, ‘Do not know’, or ‘Prefer not to answer’, with further sub-categories under each option. Participants were included for our analyses if they self-identified as ‘White’, or as ‘Asian or Asian British’ and subsequently as ‘Indian’, hereafter referred to as ‘white British’ and ‘British Indian’. The white British population was included as it made up the majority of the participants in UK Biobank, while the British Indian population was included because of the large proportion of vegetarians in this population group (25% compared to less than 2% in the overall population). The number of vegetarians in the other ethnic groups was too small to allow valid comparisons by diet groups, and therefore people of other ethnic groups were not included in these analyses.

### Diet group classification

Participants were classified into diet groups based on self-reported dietary data from the touchscreen questionnaire completed at recruitment. Participants were asked to report their frequency of consumption of processed meat (including processed poultry), unprocessed red meat (beef, lamb or mutton, pork), unprocessed poultry (such as chicken or turkey), oily fish, and other types of fish, with the possible responses ranging from “never” to “once or more daily”. Participants were also asked whether they never consumed dairy and eggs or foods containing eggs. Based on their responses to these questions, the white British participants were classified into six diet groups: regular meat eaters (red and processed meat consumption >3 times per week), low meat eaters (red and processed meat consumption ≤3 times per week), poultry eaters (participants who ate poultry but no red and processed meat), fish eaters (participants who ate fish, but not red and processed meat, or poultry), vegetarians (participants who did not eat any meat or fish), and vegans (participants who did not eat any meat, fish, dairy products or eggs). The British Indian participants were classified into two diet groups: meat eaters (ate any combination of red or processed meat or poultry) and vegetarians; the numbers of fish eaters and vegans were low in the British Indians (around 30 participants combined), and thus were not included in these analyses.

### Assay for plasma proteins

Non-fasting blood samples were collected from all UK Biobank participants by trained personnel (either a phlebotomist or a nurse) except in a small proportion (0.3%) of participants who declined, were deemed unable to, or where the attempt was abandoned for technical or health reasons. Proteomic profiling was conducted using the Olink Proximity Extension Assay on blood plasma samples in 54,219 participants, including 46,595 participants randomly selected at baseline, 6,376 participants selected by the UK Biobank Pharma Proteomics Project (UKB-PPP) consortium members at baseline and 1,268 individuals who participated in the COVID-19 repeat-imaging study (20 of whom are also in the UKB-PPP subset) [7]. The samples were shipped on dry ice to Olink Analyses Service, Uppsala, Sweden for analysis; details of the selection procedures and the technical details of the proteomics assays have been described previously [7]. In brief, the Olink Explore 3072 platform used in this study is an antibody-based assay which measures the relative abundance of 2,941 protein analytes, including 2,923 unique proteins, distributed across eight 384-plex panels (cardiometabolic, cardiometabolic II, inflammation, inflammation II, neurology, neurology II, oncology and oncology II). Measurements of proteins are expressed as normalized protein expression (NPX) values which are log-base-2 transformed. Protein values below the limit of detection (LOD) were replaced with the LOD divided by the square root of 2 [8], and protein concentrations were subsequently inverse rank normal transformed. As a result of the processing of protein data, the unit differences in protein concentrations may be interpreted as SD differences. For these analyses, we excluded three proteins with a high proportion of missing data, including GLIPR1 (99% missing), NPML (74% missing) and PCOLCE (64% missing), and thus the analyses included 2920 unique proteins.

### Inclusion and exclusion criteria

Of the 54,219 participants selected for proteomic profiling, 53,020 participants remained after the quality control procedures. Participants were further excluded if they were not of white British or British Indian ethnicity (n=2989), could not be classified into one of the pre-specified diet groups or had missing information for fasting time (n=416). After the exclusions, 49,615 participants (49,057 white British and 558 British Indian) were included in the analyses. A participant flow chart of the inclusion criteria is shown in **Supplementary figure 1**.

### Statistical analyses

Baseline characteristics of UK Biobank participants included in these analyses were tabulated by six diet groups in the white British population and by two diet groups in the British Indian population, as mean (SD) or number (%). We used multivariable adjusted linear regressions to estimate differences in protein concentrations by diet group, separately by ethnicity, using regular meat eaters as a reference group in the white British participants, and meat eaters as a reference group in the British Indians. The model was adjusted for age at recruitment (5 year categories), sex, region (London, North-West England, North-East England, Yorkshire, West Midlands, East Midlands, South-East England, South-West England, Wales, Scotland), fasting status (0–1, 2, 3, 4, 5, 6–7, ≥8 hours), body mass index (BMI; <20, 20.0–22.4, 22.5–24.9, 25.0–27.4, 27.5–29.9, 30.0–32.4, 32.5–34.9, ≥35 kg/m^2^, unknown), alcohol consumption (<1, 1–7, 8–15, ≥16 g/d, unknown), smoking status (never, previous, current <15 cigarettes/day, current ≥15 cigarettes/d, unknown) and physical activity (<10, 10-49.9, ≥50 excess metabolic equivalent of task, hr/wk, unknown). Wald tests were used to assess overall heterogeneity between diet group in each ethnicity, and for pairwise comparisons of each of the other diet groups compared to the reference group (regular meat eaters in white British participants/meat eaters in British Indians). Heterogeneity between vegetarians and vegans in the white British population was assessed based on post-estimation linear combinations of parameters for all proteins.

To account for multiple testing while considering the high correlations between the circulating proteins [9], we conducted a principal component analysis of the circulating proteins in the analysis dataset, and determined that the first 1346 principal components explained 95% of the total variation in the exposure data. Consequently, the effective number of independent tests was determined to be 1346, and the *p*-value for statistical significance was set to be 0.05/1346 = 0.000037. For the top proteins identified (the top 10 significant proteins in each pairwise comparison against the reference group, based on ranking of *p*-values), we conducted additional sensitivity analyses to evaluate the extent to which the associations may be influenced by key covariates including BMI, smoking and alcohol consumption, by presenting models with and without adjustment of these variables. Additionally, we also conducted sensitivity analyses restricted to people who self-reported to be in good and excellent health. All analyses were performed using Stata version 18.1 (StataCorp, TX, USA). All figures were generated using R version 4.2.1, the forest plots using “Jasper makes plots” package version 2-266 [10].

## Results

### Baseline characteristics

The baseline characteristics of the study population are shown by ethnicity and diet group in Table 1. Compared with white British regular meat eaters, the non-meat eaters were on average slightly younger and more likely to be women. They had lower BMI on average, reported lower alcohol consumption, more likely to have never smoked, and reported more physical activity. Compared with British Indian meat eaters, the vegetarians were more likely to be women and reported lower alcohol consumption, were more likely to have never smoked and reported lower physical activity, but BMI was not noticeably different between the two groups. Fasting time was not meaningfully different by diet group in both ethnicities.

**Table 1:** Baseline characteristics of white British and British Indian participants by diet groups in UK Biobank.

|  | White British participants |  |  |  |  |  | British Indian participants |  |
| --- | --- | --- | --- | --- | --- | --- | --- | --- |
|  | Regular meat eaters<br>N=23,243 | Low meat eaters<br>N=23,472 | Poultry eaters<br>N=486 | Fish eaters<br>N=1,081 | Vegetarians<br>N=721 | Vegans<br>N=54 | Meat eaters<br>N=391 | Vegetarians<br>N=167 |
| Age at recruitment | 57.2 (8.2) | 57.2 (8.0) | 56.6 (8.1) | 54.4 (8.2) | 52.9 (8.1) | 55.0 (7.8) | 53.9 (8.6) | 54.6 (7.8) |
| Women | 9,852<br>(42.4%) | 14,962<br>(63.7%) | 390 (80.2%) | 762 (70.5%) | 483 (67.0%) | 28 (51.9%) | 171 (43.7%) | 105 (62.9%) |
| Men | 13,391<br>(57.6%) | 8,510<br>(36.3%) | 96 (19.8%) | 319 (29.5%) | 238 (33.0%) | 26 (48.1%) | 220 (56.3%) | 62 (37.1%) |
| Fasting time (hours) | 3.8 (2.5) | 3.7 (2.3) | 3.8 (2.2) | 3.6 (2.3) | 3.7 (2.4) | 3.8 (3.0) | 4.1 (2.4) | 4.1 (2.5) |
| Body mass index (kg/m<sup>2</sup>) | 28.0 (4.8) | 27.1 (4.7) | 25.3 (4.6) | 25.3 (4.2) | 25.3 (4.4) | 25.2 (6.3) | 27.1 (4.5) | 27.0 (4.5) |
| Alcohol consumption <1g/d | 3,618<br>(15.6%) | 4,573<br>(19.5%) | 152 (31.3%) | 255 (23.6%) | 171 (23.7%) | 27 (50.0%) | 157 (40.2%) | 143 (85.6%) |
| Never smoked | 11,785<br>(50.7%) | 13,015<br>(55.4%) | 269 (55.3%) | 603 (55.8%) | 421 (58.4%) | 29 (53.7%) | 291 (74.4%) | 152 (91.0%) |
| Physical activity (MET hrs/wk) | 34.9 (38.6) | 33.8 (34.6) | 41.5 (38.3) | 37.9 (38.8) | 34.5 (32.8) | 38.9 (41.7) | 29.7 (35.8) | 22.2 (28.9) |

Numbers shown are mean (SD) or number (%).

### Differences in plasma proteins by diet group

The plasma proteins that were significantly different in white British vegetarians and vegans compared with regular meat eaters are shown as volcano plots in **Figure 1**. Significant differences in proteins in white British low meat eaters, poultry eaters and fish eaters compared with regular meat eaters, and in British Indian vegetarians compared with meat eaters are shown as volcano plots in **Supplementary figures 2-6**. Overall, 920 plasma proteins were significantly different by diet group (based on p-heterogeneity < 0.000037) in the white British participants, including 632 plasma proteins that were significantly different in one or more pairwise comparisons between regular meat eaters and at least one of the other diet groups. This includes 333 proteins (27 higher, 306 lower) in low meat eaters compared with regular meat eaters, 52 (9 higher, 43 lower) in poultry eaters, 193 (135 higher, 58 lower) in fish eaters, 266 (235 higher, 31 lower) in vegetarians and 15 (13 higher, 2 lower) in vegans; and 7 proteins were significantly different between vegetarians and vegans.

**Figure 1:**
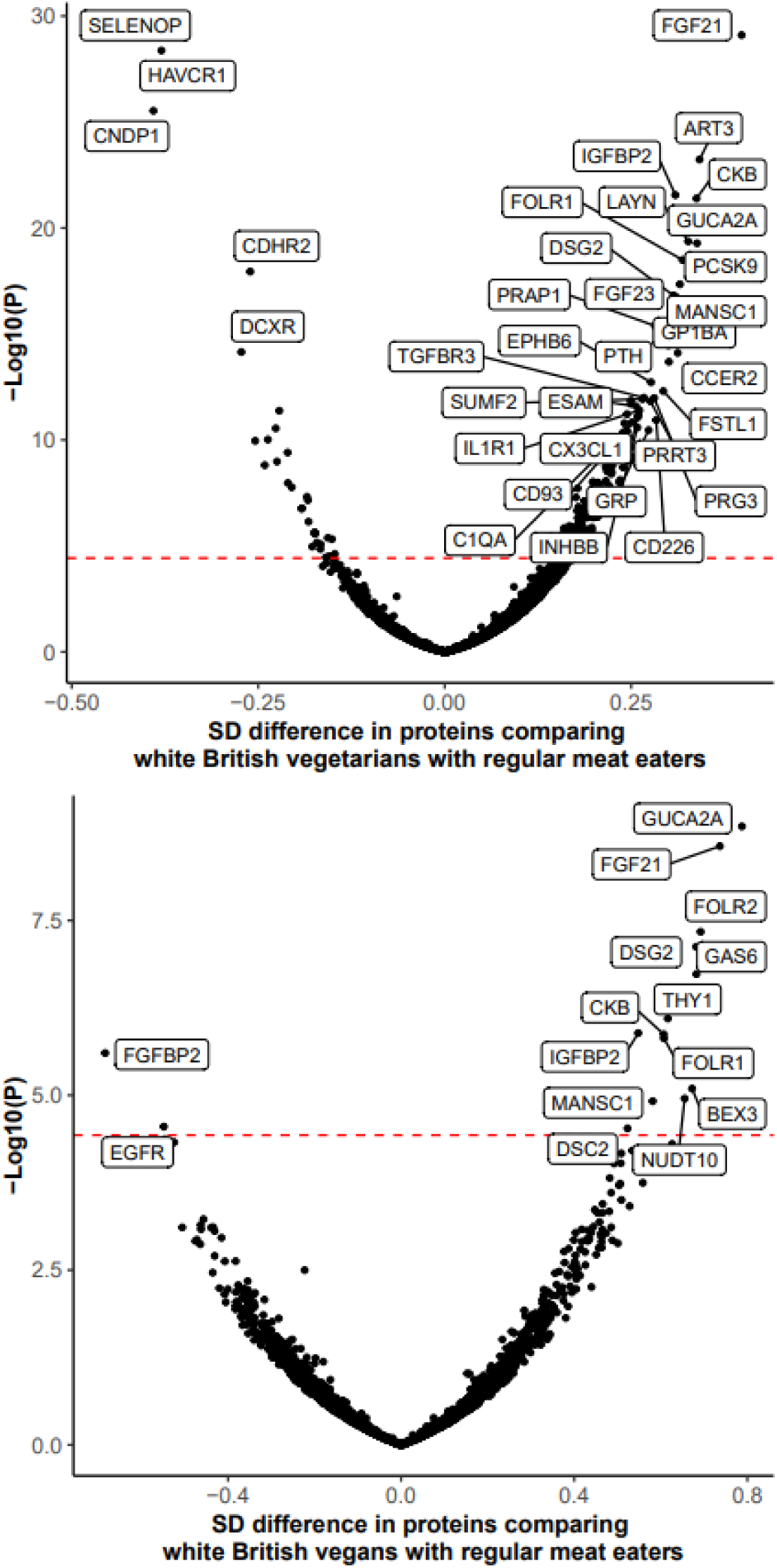
Volcano plots of proteins in white British vegetarians (top) and vegans (bottom) compared with regular meat eaters. The red dotted line signifies *p*-value threshold for statistical significance. Each dot represents one protein, which were colour-coded by whether the protein is majority expressed (>50%) in one tissue type. Results were based on the multivariable model adjusted for age at recruitment, sex, region, fasting status, body mass index, alcohol consumption, smoking status and physical activity.

In the British Indian population, only 2 proteins (SUMF2 and ATRAID) were significantly higher in vegetarians when compared with meat eaters, likely owing to the much smaller number of British Indians in the study. However, the differences in most proteins were directly consistent between the two ethnicities, as illustrated by the forest plots of top proteins by diet group and ethnicity, with the six diet groups in the white British participants and the two diet groups in British Indians (**Figures 2-3, Supplementary figures 7-9**). The complete results of all proteins are shown in **Supplementary table 1** (as excel file Supp table 1.xlsx).

**Figure 2:**
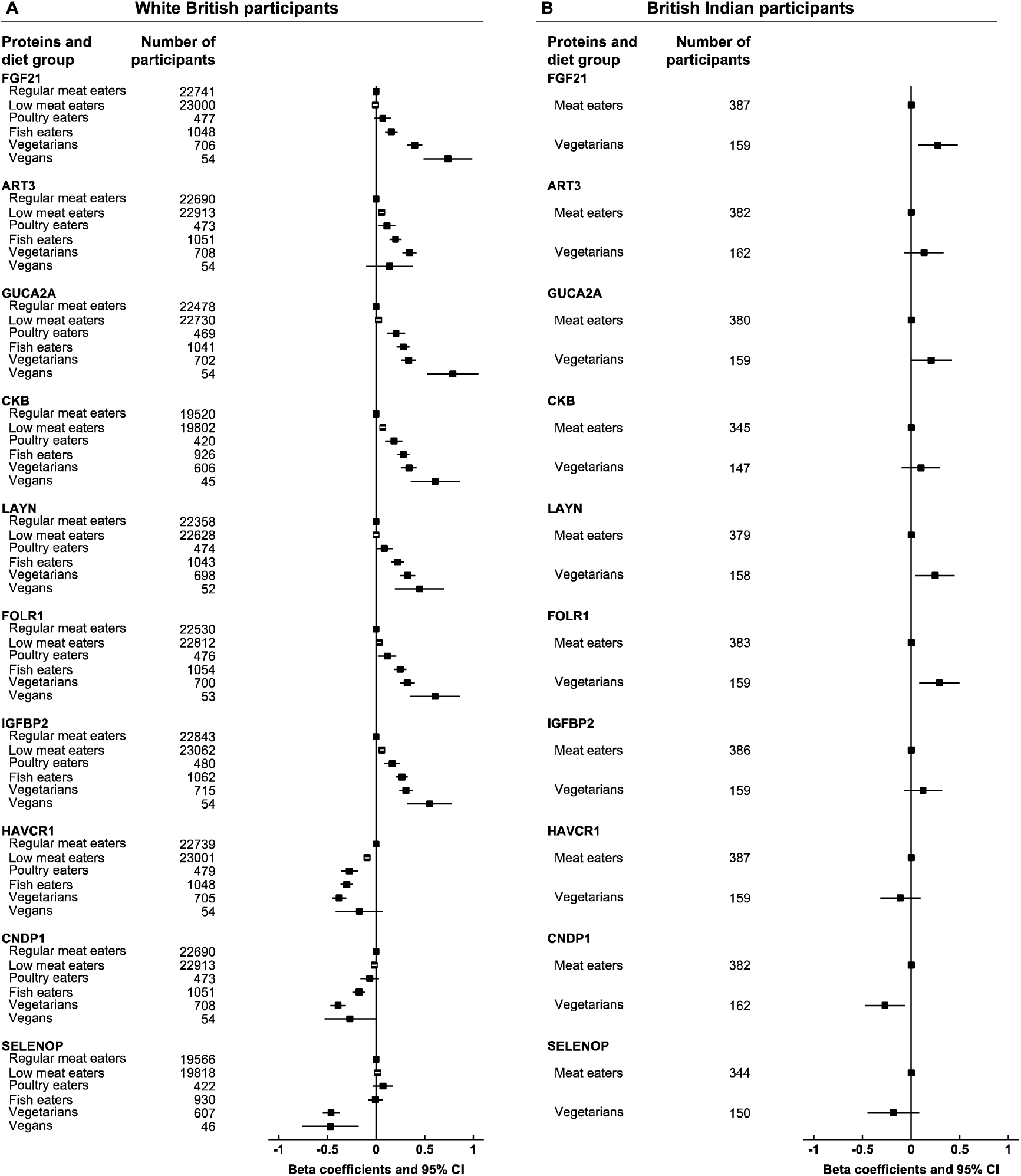
Top 10 proteins in vegetarians by diet group and ethnicity. The top 10 proteins were selected by ranking the *p*-values of proteins comparing white British vegetarians with regular meat eaters, and sorted by betas in white British vegetarians, where the betas represent SD differences. Results were based on the multivariable model adjusted for age at recruitment, sex, region, fasting status, body mass index, alcohol consumption, smoking status and physical activity.

**Figure 3:**
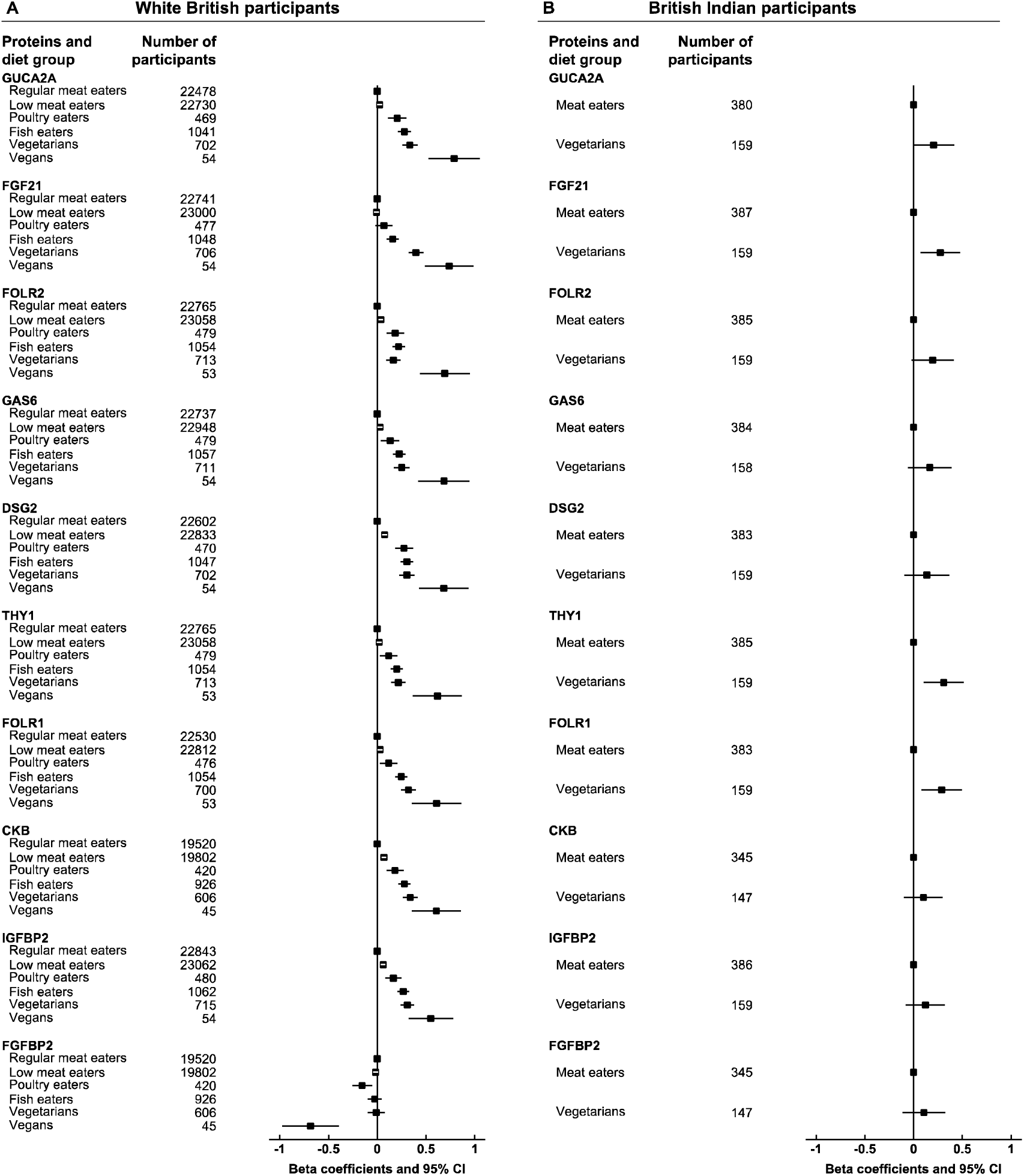
Top 10 proteins in vegans by diet group and ethnicity. The top 10 proteins were selected by ranking the *p*-values of proteins comparing white British vegans with regular meat eaters, and sorted by betas in white British vegans, where the betas represent SD differences. Results were based on the multivariable model adjusted for age at recruitment, sex, region, fasting status, body mass index, alcohol consumption, smoking status and physical activity. The results for GUCA2A, FGF21, FOLR1, CKB and IGFBP2 are the same as shown in Figure 2.

Generally, in the multivariable-adjusted model, many proteins showed a gradient effect in magnitude of differences across diet groups by degree of animal food exclusion, from regular meat eaters to vegans. Figure 2 shows the top 10 proteins in white British vegetarians (based on the ranking of pairwise p-values in vegetarians compared with regular meat eaters), and Figure 3 shows the top 10 proteins in white British vegans. Of the top proteins in vegetarians (Figure 2), compared with regular meat eaters, vegetarians and vegans both had significantly higher FGF21 (+0.40 SD in vegetarians; +0.74 SD in vegans), GUCA2A (+0.33; +0.79), CKB (+0.34, 0.60) and IGFBP2 (+0.31; +0.55) after correction for multiple testing, whereas vegetarians but not vegans had significantly higher ART3 (+0.34), LAYN (+0.32), and lower SELENOP (-0.46), CNDP1 (-0.39) and HAVCR1 (-0.38), though the differences in vegans were directionally similar. In addition to the aforementioned proteins, vegans also had substantially higher FOLR2 (+0.69), GAS6 (+0.68), DSG2 (+0.68), THY1 (+0.62), FOLR1 (+0.61) than regular meat eaters (Figure 3); the differences in these proteins were directionally consistent but less extreme in the vegetarians. Vegans, but not the other diet groups, had significantly lower FGFBP2 (-0.68) compared with regular meat eaters.

The top 10 proteins in white British low meat eaters, poultry eaters and fish eaters compared with regular meat eaters are shown in Supplementary figures 8, 9 and 10 respectively. Of these top proteins, none was uniquely different in one diet group only when compared with regular meat eaters. Similar to the observations in vegetarians, all three groups had substantially higher DSG2 and lower HAVCR1 compared with regular meat eaters; in both low meat eaters and poultry eaters HAVCR1 is the protein that exhibited the largest magnitude in difference across all proteins, while in fish eaters FGF23 showed the biggest difference. In addition, low meat eaters and poultry eaters both had lower OSM and CD99L2, the latter of which was higher in vegetarians. In sensitivity analyses of the top proteins with different levels of covariate adjustment and limited to people of good and excellent health, results were similar (**Supplementary table 2,** as excel file Supp table 2.xlsx).

## Discussion

### Summary of main findings

In this large British cohort, we observed differences in the levels of many proteins by diet group. This is the first large and comprehensive study on vegetarian diet groups and plasma proteomics, and therefore many of the substantial differences reported here are novel. In particular, we saw substantial differences in some proteins potentially associated with disease states such as HAVCR1 and DSG2 (associated with kidney disease [5,11] and inflammatory bowel disease, IBD [12], respectively), as well as proteins associated with nutritional status including IGFBP2 [13,14], and possibly FGF21 [15] and the folate receptors FOLR1 and FOLR2 [16]. The magnitudes of differences in many proteins showed a gradient effect across the diet groups, and were directionally consistent between the white British and British Indian populations.

### Comparison with previous studies

No previous studies were found that examined plasma proteomics in habitual vegetarians and vegans. A small trial of 20 participants reported minimal differences in 1306 proteins after being on a vegan diet for two weeks [17], but these results based on temporary vegan diets are not comparable to our findings among habitual vegans and vegetarians. Another study reported 837 proteins associated with different plant-based diet indices in a predominantly non-vegetarian population [18], while other previous studies have reported on proteomic profiles by other dietary patterns, including the Dietary Approaches to Stop Hypertension (DASH) diet, the Healthy Eating Index and the Mediterranean diet, as well as other data derived dietary patterns [19–22]. Whilst vegetarian and vegan diets share some common features with these other healthy dietary patterns, such as higher intakes of fruit, vegetables and whole grains, the diets are also inherently different in many other aspects. For example, while both the DASH diet and Mediterranean diet recommend reducing red and processed meat consumption, they generally do not pose any restrictions on consumption of fish and fish products, or dairy products (particularly low-fat versions) [23,24], and participants with the highest adherence to the plant-based diet index still regularly consumed animal products [25]. In contrast, vegetarian and vegan diets are defined by the complete exclusion of animal food groups. Taking into account these differences in diet as well as differences in the proteomics platforms used, it is not surprising that we have not found substantial similarities in the results; however, the previous studies also reported that people with higher adherence to the DASH diet had higher FOLR2 [19] and DSG2 [20], while those with higher adherence to the healthful plant-based diet index also had higher IGFBP2 [18],. There are only a limited number of studies examining vegetarian and vegan diets with individual proteins that were also of particular interest in this study, which are described below.

### SELENOP

Two previous small studies including 36 vegans [26] and 54 vegetarians/vegans [27] both found lower circulating levels of SELENOP in vegans/vegetarians than omnivores. SELENOP is a transporter protein for serum selenium, and previous studies have shown that vegetarians and vegans both had lower intakes of selenium compared with omnivores who include meat and fish in their diets [28]. Therefore, lower levels of SELENOP in response to lower selenium intakes in these diet groups may be expected. However, the soil type in which crops were grown has been shown to influence both selenium intake and status, thus the specific selenium content of plant foods may vary by their production region [29]. Some previous observational studies have suggested that lower circulating selenium might be associated with higher risks of some types of cancers [30,31] as well as lower bone mineral density [32,33], though a small selenium supplementation trial in the UK (39 or fewer participants in each arm) reported no clinically relevant differences in bone mineral density after 26 weeks [34].

### FGF21

In support of our findings, higher concentrations of FGF21 in vegetarians and vegans have been shown previously in a small study of 36 each of omnivores, vegetarians and vegans [35]. FGF21 has been hypothesised to be high in vegans as a downstream effect of low intake of methionine [15], an essential amino acid that is known to be limited in vegan diets [3]. Studies of FGF21 administration in rodents have found favourable effects on adiposity, lipid profiles, and non-alcohol fatty liver disease, but possible adverse effects on bone homeostasis [36]; randomised trials of FGF21 analogues also support the reduction in triglycerides [37] and improvement of fibrosis in patients with non-alcoholic steatohepatitis [38]. Consistent with these observations, we have previously reported lower BMI and body fat [39], more favourable lipid profiles [40] and lower heart disease risk [41], but also lower bone mineral density [39] and higher fracture risk [42] in vegetarians and vegans when compared with meat eaters; another study has reported lower odds of non-alcohol fatty liver disease in vegetarians than non-vegetarians [43]. Based on these differences in FGF21, as well the higher FGF23 in fish eaters and vegetarians, and lower FGFBP2 in vegans but not the other diet groups, the potential relevance of FGF signalling pathways in relation to both diet and disease risk deserve further in-depth investigation.

### GUCA2A

GUCA2A was substantially higher in all diet groups compared to regular meat eaters, with the exception of low meat eaters. GUCA2A activates the Guanylate Cyclase C receptor, which has a role in the maintenance of gut physiology including the increase of water movement into the intestinal lumen [44]. Consistent with the observations in this study, a previous EPIC-Oxford study reported a higher frequency of bowel movements in vegetarians and vegans [45]. Previous studies have found that fasting plasma levels of GUCA2A are significantly lower in patients who have been diagnosed with Crohn’s disease compared with healthy controls [46], and that loss of GUCA2A expression may be an important determinant of colorectal cancer via silencing of the GUCY2C tumour suppressor [47].

### FOLR1 and FOLR2

We found both FOLR1 and FOLR2 to be higher in non-meat eaters, with vegans having particularly high levels. FOLR1 and FOLR2 are both folate receptors which bind to and import folic acid into cells, and are usually downregulated with repletion of folate, however, this process may also be mediated by homocysteine in a positive direction [16]. While vegetarians and vegans typically have high dietary and serum folate, they may also have high homocysteine due to low vitamin B12 intakes [48,49], but no other studies have reported on folate receptor expression in vegetarians and vegans for comparison. Overexpression of folate receptors has been reported in patients with some types of cancers [50,51], and previous prospective analyses in UK Biobank have also found a positive association of both FOLR1 and FOLR2 with kidney cancer [5]. However, previous studies have not shown a higher risk of kidney cancer in vegetarians or vegans compared with meat eaters [52], and thus it is possible the upregulation of folate receptors in these groups mainly reflect differences in diet and homocysteine levels.

### IGFBP2

Similar to our findings, a previous study in EPIC-Oxford has found that compared with meat-eaters, both vegetarians and vegans had higher concentrations of IGFBP2, a binding protein for IGF-1 [13]. The mechanism for higher IGFBP2 in vegetarians and vegans is not well established but has been suggested to be related to lower dietary intakes of protein (especially from dairy) [14], essential amino acids [13], or lower total energy intake [53], which are all characteristic of vegetarian and vegan diets [3,28,39]. Insulin-like growth factors and their associated binding proteins, including IGFBP2, are involved in the regulation of cell proliferation, differentiation, and apoptosis [54,55]. However, previous studies, including from UK Biobank, have not clearly established an independent association of IGFBP2 with different cancer types [5,56–58].

### CKB

CKB or Creatine kinase B-type, is a class of creatine kinases which are cytoplasmic enzymes involved in energy homeostasis, and often elevated in cases of musculoskeletal trauma or myocardial injury [59]. CKB is the main isoform of creatine kinase in the brain, and plays a critical role in the regulation of ATP levels in neural cells; previous studies have shown that CKB levels are reduced in brain regions affected by neurodegenerative diseases such as Alzheimer’s disease and dementia [60,61]. On the other hand, elevated levels of CKB have also been reported in patients with acute brain injury or stroke [62,63], or failure or suppression of osteoclasts [64]. Given these potentially contradictory reports, the implications and relevance of higher CKB in non-meat eaters compared with meat eaters require further investigation.

### DSG2

Low and non-meat eaters, particularly vegans, had higher DSG2 in the current study. DSG2 belongs to the family of desmosomal cadherins, which includes both desmogleins (DSGs) and desmocollins (DSCs), with DSG2 and DSC2 being the major isoforms in humans, and the only isoforms expressed in the intestinal epithelium [12]. Consistently, the current study also showed that DSC2 was higher in non-meat eaters particularly vegans. Though the two proteins are structurally and functionally similar, laboratory evidence in mouse models suggest that DSG2 may have a more prominent role in the maintenance of the integrity of the intestinal epithelial barrier than DSC2 [65]. In humans, several small studies on IBD patients have found lower intestinal protein levels of DSG2 in IBD patients, suggesting a role of the protein in IBD pathogenesis [12].

### HAVCR1

HAVCR1, also known as KIM-1, has been recognised as a biomarker of kidney injury and possibly an early diagnostic marker of kidney disease [11], as well as being associated with kidney cancer [5,66]. In mice, high-fat diets or the saturated fatty acid palmitate have been shown to upregulate expression of HAVCR1 in the proximal tubules of the kidney [67–69]. In humans, vegetarians and vegans tend to have lower intake of saturated fat, which is consistent with the observation of lower HAVCR1 in the non-meat eaters (though not significantly so in vegans) in the current study. Additionally, high protein diets, especially diets high in protein from animal sources, have been suggested to have adverse effects on kidney health [70]. A previous study suggested a lower risk of chronic kidney disease in vegetarians after accounting for baseline risk of diabetes and hypertension [71]. Furthermore, as HAVCR1 is expressed on the surface of immune cells, it is also believed to have a role regulating immune responses via activation and proliferation of immune cells [11].

### Other proteins

The functions of many other proteins identified are not well established. CNDP1 is a dipeptidase, and one previous study reported higher CNDP1 in people on high protein diets [72], which would be consistent with our observation of lower CNDP1 in non-meat eaters (not statistically significant in vegans) who generally have lower protein intake. Several of the proteins identified such as ART3, GAS6, THY1 which were higher in vegetarians and/or vegans are involved in a range of signalling pathways and progression of tumour tissues [73–76], while other proteins including LAYN and CDHR2 are believed to have roles in cell adhesion [77,78]. AHSP, which was significantly higher in vegans than vegetarians, has a role in stabilising haemoglobin and its loss of function has been linked to inherited anaemia [79], but its relevance for nutritional anaemia is unclear. Overall, the reasons that these proteins appear to be influenced by diet and their implications require further research. As the first study on habitual vegetarian diet group and a large panel of circulating proteins, the current study focused on the biggest differences by diet groups, and many other differences were observed that could not be described in detail, but still deserve further investigation. The study demonstrates the utility of investigating circulating protein differences by diet groups; future research should measure these and other proteins using multiple technologies in relation to dietary factors, to replicate and expand on our findings.

### Strengths and limitations

The key strength of this study was that it showed for first time differences in circulating proteins between people of different habitual vegetarian diet groups in a large population, and many of the findings were completely novel. We have defined six diet groups in the white British population and two diet groups in the British Indian population, which allowed a detailed comparison by both dietary habits and ethnicity. Of the limitations, as with all observational studies, some level of self-selection bias may be present, which limits the generalisability of the findings. While we have adjusted for several important confounders, residual confounding by other dietary and non-dietary factors may still be present, though sensitivity analyses showed that results were consistent across all adjustment models. We are also unable to infer causality due to the cross-sectional nature of the study.

## Conclusions

In this large population-based cohort in the United Kingdom, many differences in circulating protein concentrations were observed between different diet groups. These proteins are involved in a range of different biological pathways and processes, including nutritional status, and functions related to gastrointestinal tract and kidney function. These differences likely reflect physiological differences between the different diet groups, and the implications of these differences for future disease risk, as well as specific diet-protein associations of relevant foods and nutrients, require further investigation.

## Data Availability

UK Biobank is an open access resource. Bona fide researchers can apply to use the UK Biobank dataset by registering and applying at http://ukbiobank.ac.uk/register-apply/

## Acknowledgements

This research has been conducted using UK Biobank Resource under application 67506.

## Author’s contributions

TYNT, TJK and RCT conceived and designed the research question. KSB conducted quality control of the proteomics data. TYNT analysed the data and wrote the first draft of the manuscript. All authors provided input on data analysis and interpretation of results, revised the manuscript critically for important intellectual content, and read and approved the final manuscript.

## Data sharing

The UK Biobank is an open-access resource. Bona fide researchers can apply to use the UK Biobank data set by registering and applying at http://www.ukbiobank.ac.uk/register-apply/.

