## Supplementary tables and figures for "The plasma proteome of plant-based diets: analyses of 2920 proteins in 49,615 people"

**Supplementary tables 1**: Differences in proteins by diet group compared with white British regular meat eaters or Indian meat eaters.

Please refer to excel file “Supp tables 1.xlsx”, which shows results for differences in 2920 proteins across six diet groups in white British (worksheet ‘white British participants’) and two diet groups in British Indians (worksheet ‘British Indian participants’).

In both worksheets, column A shows the protein name and column B shows the number of participants included in the analyses for the protein.

In ‘white British participants’, columns D to R shows the beta coefficients and 95% confidence intervals for low meat eaters, poultry eaters, fish eaters, vegetarians, and vegans compared with regular eaters; column S shows the *p*-heterogeneity across all diet groups, columns T to X shows *p*-value for pairwise comparisons of each diet group against regular meat eaters, column Y the *p*-value for pairwise comparisons of vegetarians and vegans.

In ‘British Indian participants’, columns D to F shows the beta coefficients and 95% confidence in vegetarians compared with meat eaters, and column G shows the corresponding *p*-value.

All estimates were based on multivariable model adjusted for age at recruitment, sex, region, fasting status, body mass index, alcohol consumption, smoking status and physical activity.

54,219 participants selected for proteomic profiling

Excluded 2989 participants of other (i.e. not white British or British Indian) or unknown ethnicities

Excluded 1199 participants whose protein measurements did not pass quality control checks

53,020 participants

Excluded 416 participants who could not be classified into one of the prespecified diet groups or with missing information on fasting time

49,615 participants, including
49,057 white British and
558 British Indian

**Supplementary figure 1**: Participant flow chart of the study.

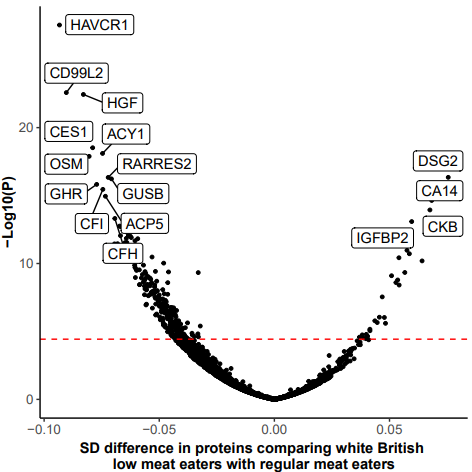

**Supplementary figure 2**: Volcano plots of proteins in white British low meat eaters compared with regular meat eaters.

The red dotted line signifies *p*-value threshold for statistical significance. Each dot represents one protein, which were colour-coded by whether the protein is majority expressed (>50%) in one tissue type. Results were based on the multivariable model adjusted for age at recruitment, sex, region, fasting status, body mass index, alcohol consumption, smoking status and physical activity.

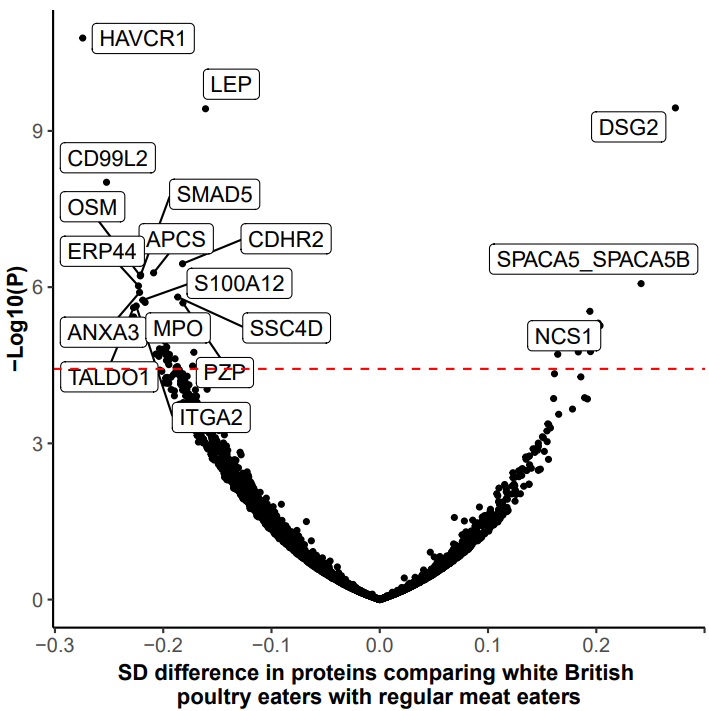

**Supplementary figure 3**: Volcano plots of proteins in white British poultry eaters compared with regular meat eaters.

The red dotted line signifies *p*-value threshold for statistical significance. Each dot represents one protein, which were colour-coded by whether the protein is majority expressed (>50%) in one tissue type. Results were based on the multivariable model adjusted for age at recruitment, sex, region, fasting status, body mass index, alcohol consumption, smoking status and physical activity.

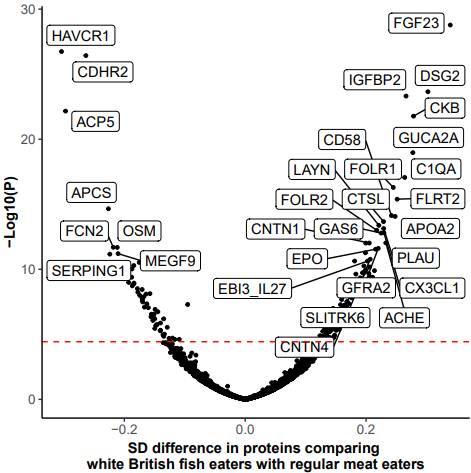

**Supplementary figure 4**: Volcano plots of proteins in white British fish eaters compared with regular meat eaters.

The red dotted line signifies *p*-value threshold for statistical significance. Each dot represents one protein, which were colour-coded by whether the protein is majority expressed (>50%) in one tissue type. Results were based on the multivariable model adjusted for age at recruitment, sex, region, fasting status, body mass index, alcohol consumption, smoking status and physical activity.

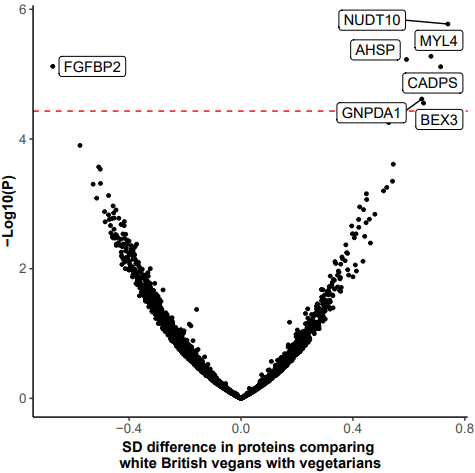

**Supplementary figure 5**: Volcano plots of proteins in white British vegans compared with vegetarians.

The red dotted line signifies *p*-value threshold for statistical significance. Each dot represents one protein, which were colour-coded by whether the protein is majority expressed (>50%) in one tissue type. Results were based on the multivariable model adjusted for age at recruitment, sex, region, fasting status, body mass index, alcohol consumption, smoking status and physical activity.

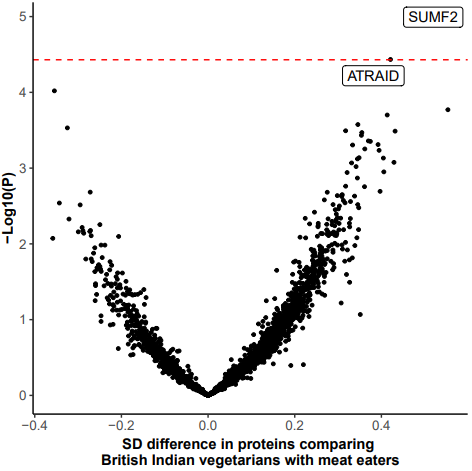

**Supplementary figure 6**: Volcano plots of proteins in British Indian vegetarians compared with meat eaters.

The red dotted line signifies *p*-value threshold for statistical significance. Each dot represents one protein, which were colour-coded by whether the protein is majority expressed (>50%) in one tissue type. Results were based on the multivariable model adjusted for age at recruitment, sex, region, fasting status, body mass index, alcohol consumption, smoking status and physical activity.

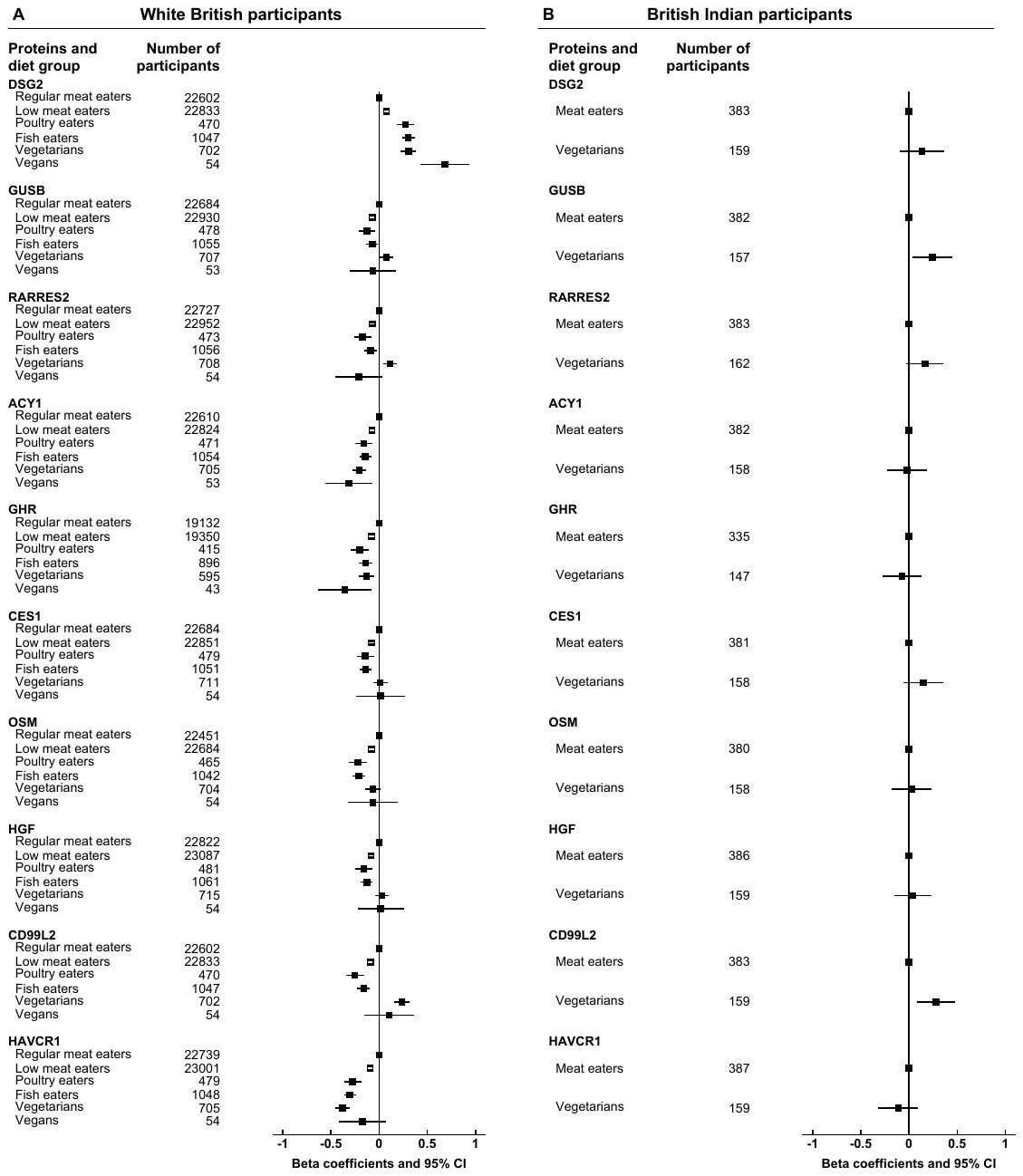

**Supplementary figure 7**: Top 10 proteins in low meat eaters by diet group and ethnicity.

The top 10 proteins were selected by ranking the *p*-values of proteins comparing white British low meat eaters with regular meat eaters, and sorted by betas in white British low meat eaters, where the betas represent SD differences. Results were based on the multivariable model adjusted for age at recruitment, sex, region, fasting status, body mass index, alcohol consumption, smoking status and physical activity.

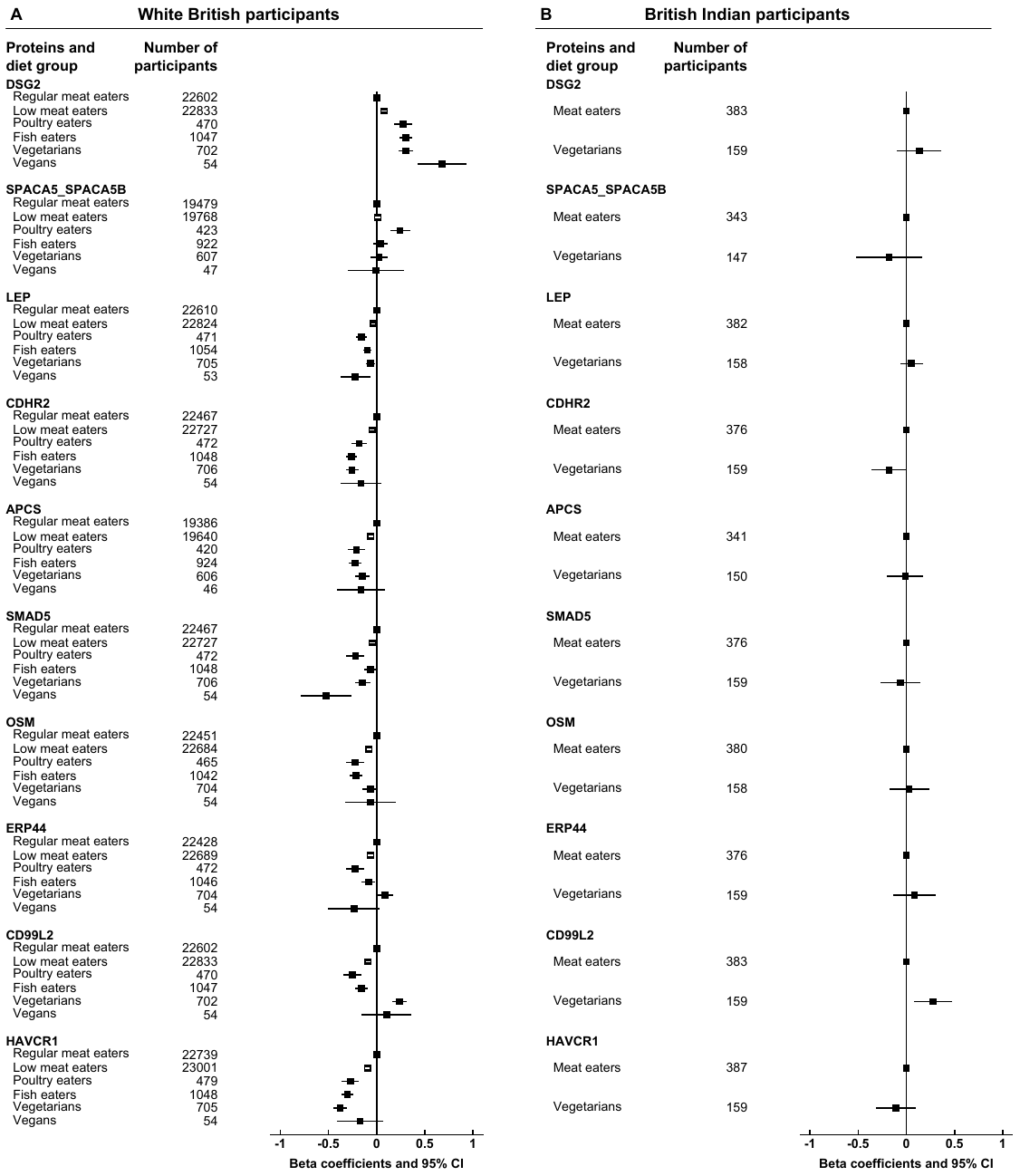

**Supplementary figure 8**: Top 10 proteins in poultry eaters by diet group and ethnicity.

The top 10 proteins were selected by ranking the *p*-values of proteins comparing white British poultry eaters with regular meat eaters, and sorted by betas in white British poultry eaters, where the betas represent SD differences. Results were based on the multivariable model adjusted for age at recruitment, sex, region, fasting status, body mass index, alcohol consumption, smoking status and physical activity.

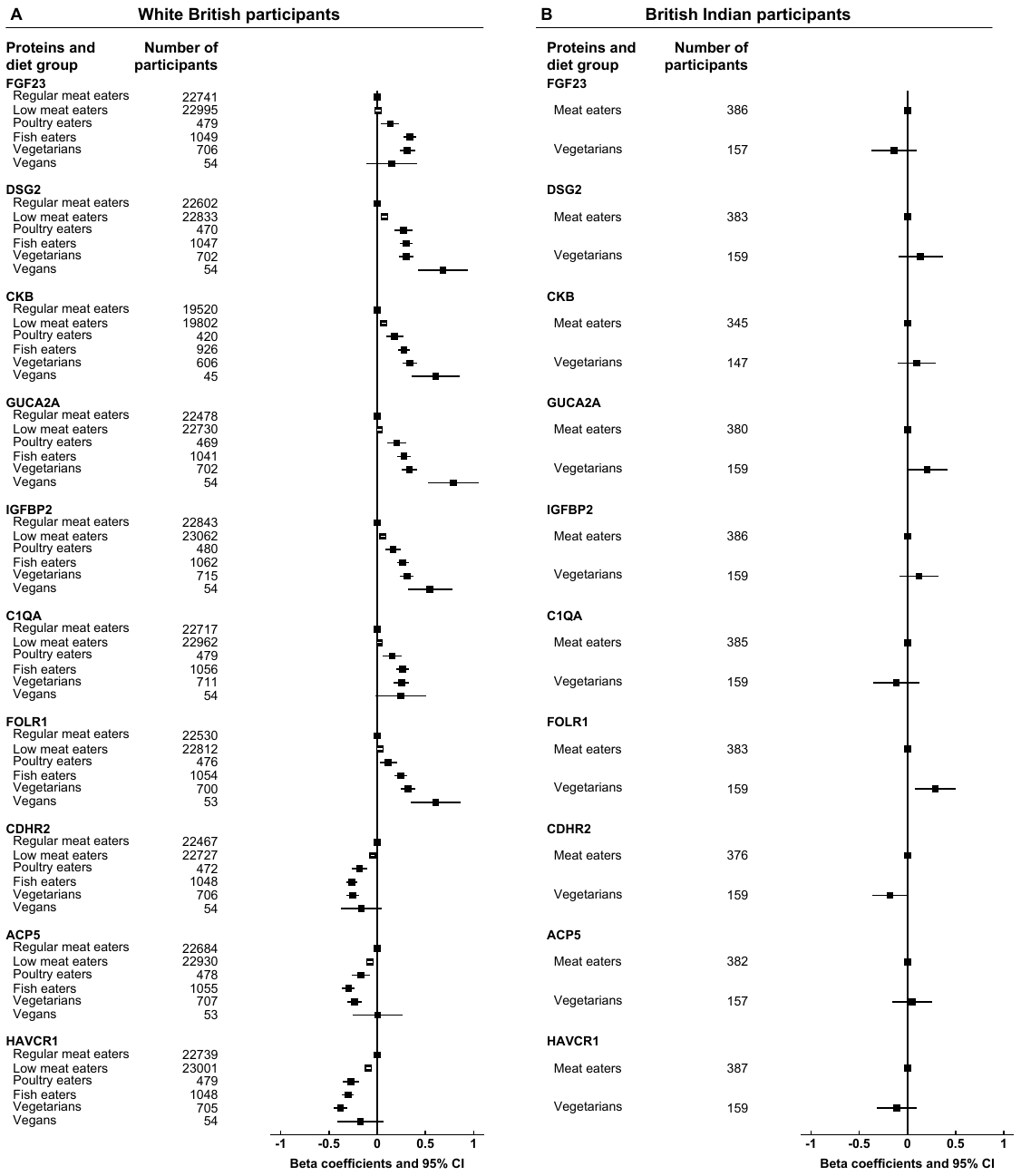

**Supplementary figure 9**: Top 10 proteins in fish eaters by diet group and ethnicity.

The top 10 proteins were selected by ranking the *p*-values of proteins comparing white British fish eaters with regular meat eaters, and sorted by betas in white British fish eaters, where the betas represent SD differences. Results were based on the multivariable model adjusted for age at recruitment, sex, region, fasting status, body mass index, alcohol consumption, smoking status and physical activity.

**Supplementary tables 2**: Sensitivity analyses of top proteins with different covariate adjustment and restricted to people who reported good or excellent health.

Please refer to excel file “Supp tables 2.xlsx”, which shows results for differences in selected proteins across six diet groups in white British participants and two diet groups in British participants. All worksheets are named as ‘white_*protein_name*’ or ‘indian_*protein_name*’, which correspond to results across different adjustment models for the protein (as indicated in *protein_name*) in white British and British Indian respectively.

In all worksheets, column A shows the adjustment model and column B shows the number of participants included in the analyses for the protein. ‘Main model’ (row 2) refers to the multivariable model adjusted for age at recruitment, sex, region, fasting status, body mass index, alcohol consumption, smoking status and physical activity. ‘Minus BMI’ (row 3) refers to the multivariable model without BMI; ‘Minus smoking’ (row 4) refers to the multivariable model without smoking; ‘Minus alcohol consumption’ (row 5) refers to the multivariable model without alcohol consumption. ‘Good or excellent health’ (row 6) refers to analyses restricted to people who self-reported to be in good or excellent health, with adjustment as in the multivariable model.

In all ‘white_*protein_name*’ worksheets, columns C to Q shows the beta coefficients and 95% confidence intervals for low meat eaters, poultry eaters, fish eaters, vegetarians, and vegans compared with regular eaters; column R shows the *p*-heterogeneity across all diet groups, columns S to W shows *p*-value for pairwise comparisons of each diet group against regular meat eaters, column X the *p*-value for pairwise comparisons of vegetarians and vegans.

In all ‘indian_*protein_name*’ worksheets, columns C to E shows the beta coefficients and 95% confidence in vegetarians compared with meat eaters, and column F shows the corresponding *p*-value.
